# Effect of monitoring and evaluation data management and use on Direct Health Facility Financing implementation effectiveness in urban and rural Tanzania: translating stakeholder perceptions of the DHFF M&E framework

**DOI:** 10.64898/2026.05.09.26352491

**Authors:** Deogratias. F. Mpenzi, Deus. D. Ngaruko, Roger Myrick

## Abstract

**Background:** Tanzania’s Direct Health Facility Financing (DHFF) reform was introduced to strengthen primary health care through decentralized financing, autonomy, and accountability, but persistent weaknesses in monitoring and evaluation (M&E) data management and use continue to constrain implementation effectiveness, particularly in rural settings.

**Methods:** A convergent mixed-methods design was used to examine how M&E data management and use influence DHFF implementation effectiveness in an urban council (Kinondoni Municipal Council, KMC) and a rural council (Morogoro District Council, MDC), while also assessing the role of stakeholder perceptions of the DHFF M&E framework and contextual variation. Quantitative data were analyzed using descriptive statistics, relative importance indices, regression and ANOVA, while qualitative data from key informant interviews and focus group discussions were thematically analyzed and triangulated with quantitative results.

**Results:** Of 233 respondents analysed, 51.1% were from Morogoro District Council, 48.9% from Kinondoni Municipal Council, 51.2% worked in rural settings, 42.9% were from health centres, and 38.2% from dispensaries, providing an analytically useful spread across managerial and frontline contexts relevant to DHFF implementation. Descriptive statistics showed generally favourable perceptions across the five major constructs, with mean scores ranging from 3.09 for M&E capacity to 3.73 for urban-rural M&E practice context, while DHFF implementation effectiveness scored 3.71 overall. Data quality checks showed acceptable factor loadings above 0.4, reliability coefficients above 0.7, bivariate correlations of 0.34-0.76, and VIF values of 1.31-2.95, indicating that the dataset was screened, cleaned and analytically fit for regression and ANOVA modelling. In the aggregated model, the explanatory variables jointly accounted for about 52% of the variation in DHFF implementation effectiveness, with M&E data management and use, stakeholder perceptions of the DHFF M&E framework, and urban-rural context emerging as the most influential predictors. Qualitative testimonies clarified these patterns: one council respondent explained, “We have DHIS2… GoTHOMIS… FFARS… also PlanRep,” while another facility respondent observed, “We only add up numbers for the monthly report—we don’t really analyze what they mean,” illustrating the contrast between data availability and meaningful local use.

**Conclusions:** DHFF implementation effectiveness in Tanzania depends substantially on robust M&E data management and use, supportive stakeholder perceptions of the M&E framework, and context-sensitive strategies that address persistent urban–rural inequities. Strengthening technical capacity, digital infrastructure, participatory governance and feedback systems is essential for sustaining DHFF gains and improving equitable service delivery.

## Introduction

Direct Health Facility Financing (DHFF) is one of Tanzania’s major health financing reforms designed to transfer funds directly to primary health facilities in order to improve autonomy, accountability, efficiency, and responsiveness to local health needs (1,2,27,38,39,59). Available empirical literature indicates that DHFF has improved managerial autonomy, timeliness of resource allocation, budget execution, and selected aspects of service delivery, although these gains remain constrained by uneven implementation capacity, limited supervision, inadequate financial monitoring, and weak local use of routine data (1–4,27,38,39,62,63). Within this reform environment, the ability of facilities and councils to manage and use monitoring and evaluation (M&E) data effectively has become increasingly important for understanding whether financial decentralization is translating into better health services and stronger health-system performance (5–10,33,42,51–53,61).

Studies from Tanzania consistently show that while digital systems and interoperability initiatives have improved the availability of routine health information, actual use of data for planning, budgeting, performance review, and corrective action remains weak (5–10,33,42,61). This weakness is linked to poor data quality, limited technical capacity, weak feedback systems, infrastructure constraints, and a persistent culture of reporting for compliance rather than decision-making (6–10,13–15,33,44,55,61). Similar findings are reflected in African and global literature, where routine health information systems are recognized as necessary but insufficient unless supported by governance, interoperability, organizational learning, participatory monitoring, and stronger demand for data use in decision-making (11–23,30,40,45,47–53,65).

The empirical review further suggests that stakeholder perceptions of the DHFF M&E framework matter because they shape how actors interpret the relevance, usefulness, fairness, and legitimacy of routine monitoring arrangements (16,17,22,23,28–31,35,50,51). Tanzanian and related decentralization studies show that district managers, facility staff, and local leaders often value DHFF and associated governance arrangements for enhancing transparency, autonomy, and accountability, yet many community actors and frontline implementers remain excluded by top-down tool design, limited analytic support, and weak links between monitoring, interpretation, and remedial action (2,3,24,27,29,37,38,50,60). These concerns are particularly important in rural settings, where staff shortages, remoteness, weak connectivity, technical support delays, and lower service readiness can reduce the practical value of data systems even when formal reporting structures are in place (15,24,25,32,35–37,41,54,56,57).

Urban-rural differences are especially relevant to DHFF because facilities do not operate under uniform conditions (24,32,35–39,41,56,57,59,63). Evidence from Tanzania and the wider African literature shows that urban facilities tend to have better infrastructure, stronger staffing profiles, more reliable internet access, and more learning-oriented M&E cultures, while rural facilities are more likely to experience compliance-driven reporting, delayed technical support, weaker feedback loops, and reduced use of data for local action (5–10,24,32,33,36,42,44,56,57,59–63). These contextual differences imply that the effect of M&E data management and use on DHFF implementation effectiveness may not be uniform across settings, and they also help explain why gains in autonomy or funding flow do not automatically translate into equitable improvements in service delivery, financial protection, or primary health care performance (25,26,28,31,32,41,54,58,64).

This article is derived from the broader study that examined M&E capacity, planning and budgeting, data management and use, stakeholder perceptions of the DHFF M&E framework, and contextual practices as determinants of DHFF implementation effectiveness in Tanzania. The conceptual framing positions M&E data management and use as a core explanatory factor, while stakeholder perceptions and urban-rural context are treated as intervening conditions shaping implementation outcomes at primary health facilities. Accordingly, this article focuses specifically on the effect of M&E data management and use on DHFF implementation effectiveness, while translating the mediating role of stakeholder perceptions of the DHFF M&E framework and the differences between urban and rural contexts.

## Materials and methods

### Study design

The study adopted a convergent mixed-methods design combining quantitative and qualitative approaches to examine the influence of M&E practices on DHFF implementation effectiveness in Tanzania. This design was appropriate because it enabled concurrent assessment of measurable relationships between key variables and in-depth exploration of stakeholder experiences, meanings and contextual variations relevant to DHFF implementation.

### Study area

The research was conducted in two councils implementing DHFF: Kinondoni Municipal Council (KMC), representing an urban context, and Morogoro District Council (MDC), representing a rural context. These councils were selected to capture contextual contrasts in infrastructure, staffing, information system functionality, service demand and managerial environment, all of which were relevant to M&E data management and use.

### Study population and sampling

The study population included facility in-charges, health workers, Health Facility Governing Committee (HFGC) members and council-level managers involved in DHFF implementation and M&E activities. Quantitative respondents were drawn from sampled primary health facilities, while qualitative participants were purposively selected for their direct experience with DHFF planning, budgeting, reporting, data use and governance. The study adopting Tabachnick and Fidell ,2007 sample computation approach (N≥50+8m), obtained a sample size of 233. Its distribution is well indicated in respondents ‘demographic data table 1.

**Table 1:** Respondents’’ demographic data.

| Variable | Frequency | Percent | Cumulative Percent |
| --- | --- | --- | --- |
| **Region** |  |  |  |
| *Dar Es Salaam* | 114 | 48.93 | 48.93 |
| *Morogoro* | 119 | 51.07 | 100 |
| **Council** |  |  |  |
| *Kinondoni Municipal Council (KMC)* | 105 | 48.84 | 48.84 |
| *Mvomero District Council (MDC)* | 110 | 51.16 | 100 |
| **Location** |  |  |  |
| *Urban (KMC)* | 105 | 48.84 | 48.84 |
| *Rural (MDC)* | 110 | 51.16 | 100 |
| **Level** |  |  |  |
| *RHMT* | 18 | 7.73 | 7.73 |
| *CHMT* | 26 | 11.16 | 18.88 |
| *Health Center* | 100 | 42.92 | 61.8 |
| *Dispensary* | 89 | 38.2 | 100 |
| **Facility** |  |  |  |
| *Boko Dispensary -KMC* | 16 | 8.47 | 8.47 |
| *Bunju Health Center-KMC* | 16 | 8.47 | 16.93 |
| *Chazi Health Center-MDC* | 19 | 10.05 | 26.98 |
| *Dakawa Dispensary- MDC* | 16 | 8.47 | 35.45 |
| *Kambangwa Dispensary -KMC* | 15 | 7.94 | 43.39 |
| *Kinodoni Health Center -KMC* | 12 | 6.35 | 49.74 |
| *Lusanga Dispensary-MDC* | 15 | 7.94 | 57.67 |
| *Magomeni Dispensary -KMC* | 18 | 9.52 | 67.20 |
| *Magomeni Health Center-KMC* | 15 | 7.94 | 75.13 |
| *Melela Health Center-MDC* | 18 | 9.52 | 84.66 |
| *Mikongeni Dispensary-MDC* | 10 | 5.29 | 89.95 |
| *Mlali Health Center-MDC* | 19 | 10.05 | 100.00 |
| **Sex** |  |  |  |
| *Male* | 104 | 44.64 | 44.64 |
| *Female* | 129 | 55.36 | 100 |
| **Age** |  |  |  |
| *<30* | 16 | 6.9 | 6.9 |
| *31-40* | 75 | 32.33 | 39.22 |
| *41-50* | 100 | 43.1 | 82.33 |
| *51-60* | 39 | 16.81 | 99.14 |
| *61 and above* | 2 | 0.86 | 100 |
| **Level of Education** |  |  |  |
| *Std VII* | 35 | 15.02 | 15.02 |
| *Form IV* | 13 | 5.58 | 20.6 |
| *Form VI* | 1 | 0.43 | 21.03 |
| *Certificate* | 28 | 12.02 | 33.05 |
| *Diploma* | 88 | 37.77 | 70.82 |
| *Bachelor* | 41 | 17.6 | 88.41 |
| *Masters* | 27 | 11.59 | 100 |
| **DHFF Years of Experience** |  |  |  |
| *1 Year* | 30 | 12.88 | 12.88 |
| 2 Years | 16 | 6.87 | 19.74 |
| 3 Years | 19 | 8.15 | 27.9 |
| 4 Years | 45 | 19.31 | 47.21 |
| 5 years and above | 123 | 52.79 | 100 |
| **Respondent Type** |  |  |  |
| HFGC | 85 | 36.48 | 36.48 |
| HFS | 104 | 44.64 | 81.12 |
| CHMT | 26 | 11.16 | 92.27 |
| RHMT | 18 | 7.73 | 100 |

### Data collection

Quantitative data were collected using a structured questionnaire covering M&E capacity, M&E planning and budgeting, data management and use, stakeholder perceptions of the DHFF M&E framework, contextual practices and DHFF implementation effectiveness. Qualitative data were collected through general interviews for key informants using guides designed to explore routine data practices, experiences with DHFF, governance processes, perceived relevance of the M&E framework and contextual enablers or barriers in urban and rural facilities.

### Study variables

The principal independent variable in this article was M&E data management and use, operationalized through collection, analysis, sharing and utilization of M&E data in DHFF implementation. The dependent variable was DHFF implementation effectiveness, measured mainly through perceived changes in availability and delivery of quality health services, supported by organizational, behavioural changes and output-level improvements. Stakeholder perceptions of the DHFF M&E framework and urban–rural context were treated as intervening variables influencing how M&E data practices translated into implementation outcomes.

### Data analysis

Data were first screened and cleaned in STATA version 18 to identify entry errors, confirm correct coding and labelling, and ensure consistency of composite variables before analysis. Quantitative analysis then proceeded through descriptive statistics, crosstabulations, summary measures, relative importance indices, regression and ANOVA, while factor analysis, reliability testing, heteroscedasticity checks and multicollinearity diagnostics were used to confirm analytic fitness of the measures and models. Qualitative data from interviews were transcribed, organized into themes aligned to the study variables, and triangulated with quantitative findings so that numerical patterns could be interpreted alongside stakeholder testimonies from urban and rural settings.

### Ethical considerations

The study followed formal ethical procedures, including obtaining necessary approvals, permissions and informed consent before fieldwork commenced. Confidentiality and anonymity were maintained throughout data collection, transcription, analysis and reporting.

At Open University of Tanzania, final proposal processing procedure after approval of the final presentation panel and critical reviewers’ examinations. The proposal is set for Post graduate studies Committee for further standard and ethical qualification examination. After a successful examination, the committee grant the research clearance permission.

The University wrote/ offered a research clearance permit to all research sites Authorities (Ministry Head Quarters, Regional and District Level Authorities), introducing the researcher and the study area/focus. Then the research site authorities at all levels communicated through official letters to allow the research to collect, process and report on the data findings on the focus area.

The researcher paid courtesy visits to all research sites to introduce the study and seek participation consents. Through prior debriefings with managements and study participants clarifications were made to all queries raised, and participants agreed to proceed attending the data collection tools Research clearance permits from the Open University to all Research sites and the permissions from the later have submitted for evidence

## Results

### Respondent profile

The analysis was based on 233 respondents, with near balance by region and council: 114 respondents (48.93%) were from Dar es Salaam and 119 (51.07%) from Morogoro; 105 (48.84%) were from KMC and 110 (51.16%) from MDC. Urban respondents represented 48.84% and rural respondents 51.16%, while the largest respondent groups were health-centre staff (42.92%) and dispensary staff (38.2%), followed by CHMT (11.16%) and RHMT (7.73%). These distributions are relevant because the study sought to compare M&E practices across governance levels and across urban and rural implementation environments under DHFF.

The educational profile also has direct relevance to interpretation of the findings. Most respondents (65.2%) had diploma level or below, whereas bachelor’s and master’s degree holders were concentrated at RHMT and CHMT levels; frontline health centres and dispensaries were dominated by diploma-level and below staff. This pattern helps explain why higher-level teams showed stronger agreement on M&E variables and why lower-level facilities appeared more constrained in data analysis, planning and interpretation capacity (*Table 1*).

### Data quality and cleanliness

Before analysis, questionnaire data were screened and cleaned in STATA to ensure correct coding, labelling and entry accuracy, and composite scores were then computed from the 1-5 Likert items. Construct validation procedures indicated acceptable factor loadings above 0.4, with the example M&E-capacity factor structure showing most uniqueness values around 0.5 or lower, suggesting that the retained items were reasonably explained by the latent constructs *(Table 2)*. Reliability was also satisfactory across all main variables and outcome checks, with scale reliability coefficients above 0.7, including 0.9488 for M&E data management and use, 0.9339 for stakeholder perception, and 0.7974 for urban-rural context *(Table 3)*.

**Table 2:** Factor loading for M&E data management and use.

| Table 2: Factor loading M&E data management and use |  |  |  |  |  |  |  |
| --- | --- | --- | --- | --- | --- | --- | --- |
| Variable | Factor1 | Factor2 | Uniqueness | Variable | Factor1 | Factor2 | Uniqueness |
| d1 | 0.6060 | 0.0905 | 0.6245 | d20 | 0.6122 | -0.0343 | 0.6241 |
| d2 | 0.4555 | 0.2748 | 0.7170 | d21 | 0.4109 | 0.3288 | 0.7231 |
| d3 | 0.7135 | 0.1590 | 0.4656 | d22 | 0.4414 | 0.1555 | 0.7810 |
| d4 | 0.6996 | 0.1680 | 0.4824 | d23 | 0.7216 | -0.1742 | 0.4490 |
| d5 | 0.6160 | 0.0965 | 0.6113 | d24 | 0.6719 | -0.4442 | 0.3512 |
| d6 | 0.6847 | 0.1852 | 0.4970 | d25 | 0.7215 | -0.4315 | 0.2933 |
| d7 | 0.6092 | 0.1651 | 0.6016 | d26 | 0.5832 | -0.0150 | 0.6596 |
| d8 | 0.6811 | 0.0978 | 0.5265 | d27 | 0.6081 | -0.1749 | 0.5997 |
| d9 | 0.5431 | 0.1432 | 0.6845 | d28 | 0.6605 | -0.1961 | 0.5252 |
| d10 | 0.5466 | 0.1220 | 0.6864 | d29 | 0.2309 | -0.1522 | 0.9235 |
| d11 | 0.6987 | 0.0323 | 0.5108 | d30 | 0.5631 | -0.1244 | 0.6674 |
| d12 | 0.6944 | 0.0887 | 0.5100 | d31 | 0.3063 | 0.4718 | 0.6836 |
| d13 | 0.5809 | -0.1047 | 0.6516 | d32 | 0.3014 | 0.4673 | 0.6908 |
| d14 | 0.6947 | -0.0427 | 0.5155 | d33 | 0.4196 | 0.2730 | 0.7494 |
| d15 | 0.7233 | 0.1027 | 0.4663 | d34 | 0.5939 | -0.2138 | 0.6016 |
| d16 | 0.7118 | 0.0127 | 0.4932 | d35 | 0.7146 | -0.1393 | 0.4700 |
| d17 | 0.6515 | -0.0878 | 0.5679 | d36 | 0.7137 | -0.1186 | 0.4766 |
| d18 | 0.7403 | -0.0241 | 0.4513 | d37 | 0.7001 | -0.1980 | 0.4707 |
| d19 | 0.7521 | 0.0553 | 0.4312 |  |  |  |  |

**Table 3:** Reliability test for all main variables.

|  | Independent Variables |  |  |  |  | Outcome Variable | Check1 | Check 2 | Check 3 |
| --- | --- | --- | --- | --- | --- | --- | --- | --- | --- |
|  | M&E Capacity | M&E planning | M&E data management use | Stakeholders' perception on DHFF M&E Framework | ME Practices (urban Rural settings) | Availability of quality health service | Organizational changes | Behavioural changes | DHFF Outputs |
| AIC | 0.5323842 | 0.4575486 | 0.4246085 | 0.7149808 | 0.4966463 | 0.4633765 | 0.5149356 | 0.6886285 | 0.4624155 |
| NIS | 14 | 16 | 37 | 14 | 10 | 10 | 8 | 5 | 7 |
| SRC | 0.8823 | 0.8771 | 0.9488 | 0.9339 | 0.7974 | 0.9062 | 0.9094 | 0.931 | 0.8908 |
| SRC | 0.8823 | 0.8771 | 0.9488 | 0.9339 | 0.7974 | 0.9062 | 0.9094 | 0.931 | 0.8908 |

Average interitem covariance (AIC), Number of items in the scale (NIS) and Scale reliability coefficient (SRC)

Additional diagnostics supported the integrity of the analytic models. The correlation matrix showed moderate relationships among predictors, ranging from 0.34 to 0.76, remaining below the common multicollinearity alert threshold *(Table 4).* VIF values ranged from 1.31 to 2.95, with a mean VIF of 2.25, indicating no serious multicollinearity problem (*Table 5*). Measures of dispersion further showed standard deviations from 0.68 to 0.83, small-to-moderate negative skewness, and kurtosis values that were considered acceptable for the multivariate analyses used in the study (*Table 6*).

**Table 4:** Correlation Matrix all main variables.

| Table 4: Correlation matrix among all independent variables in the study (Obs=233) |  |  |  |  |  |
| --- | --- | --- | --- | --- | --- |
|  | CME | PME | CASUME | SPMEF | MEPERUC |
| mecap (cme) | 1.000 |  |  |  |  |
| meplan (pme) | 0.6733 | 1.000 |  |  |  |
| medata (casume) | 0.6332 | 0.7618 | 1.000 |  |  |
| meperce (spmef) | 0.6182 | 0.6493 | 0.6536 | 1.000 |  |
| mecontext (meperuc) | 0.3739 | 0.4382 | 0.4605 | 0.3396 | 1.000 |

**Table 5:** Variable Inflation factor all main variables.

| Table 5: Variable Inflation Factor (VIF) |  |  |
| --- | --- | --- |
| Variable | VIF | 1/VIF |
| CME | 2.95 | 0.339498 |
| PME | 2.84 | 0.352522 |
| CASUME | 2.11 | 0.474651 |
| SPMEF | 2.07 | 0.482391 |
| MEPERUC | 1.31 | 0.76615 |
| Mean VIF | 2.25 |  |

**Table 6:**
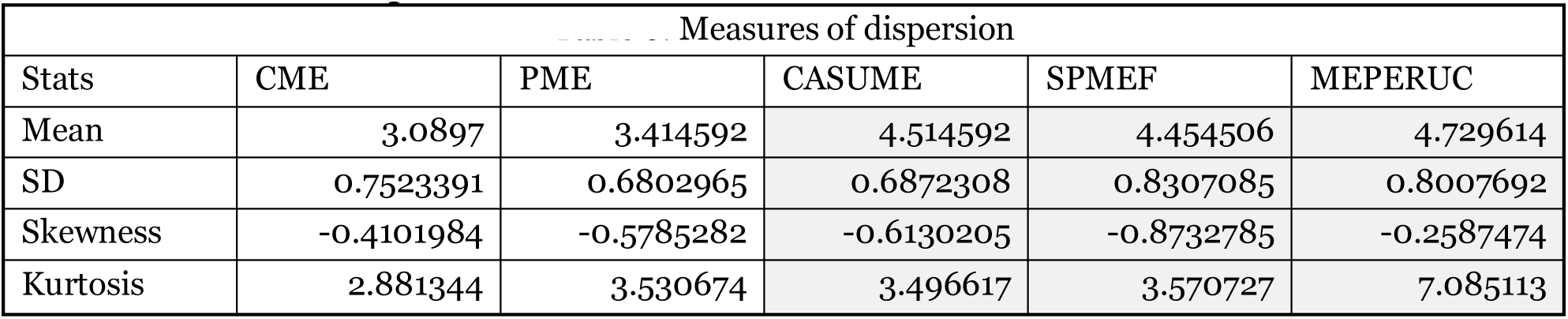
Measures of dispersion for all main variables.

| Table 6: Measures of dispersion |  |  |  |  |  |
| --- | --- | --- | --- | --- | --- |
| Stats | CME | PME | CASUME | SPMEF | MEPERUC |
| Mean | 3.0897 | 3.414592 | 4.514592 | 4.454506 | 4.729614 |
| SD | 0.7523391 | 0.6802965 | 0.6872308 | 0.8307085 | 0.8007692 |
| Skewness | -0.4101984 | -0.5785282 | -0.6130205 | -0.8732785 | -0.2587474 |
| Kurtosis | 2.881344 | 3.530674 | 3.496617 | 3.570727 | 7.085113 |

Qualitative testimonies also reflected active checking and harmonization of data at facility level. One respondent reported, “*Despite limited capacity we cross-check the data and see if they are OK or some issues for discussion, harmonization, or immediate attention*,” while another said, “*In clinical meetings we have a tendency of briefing each other on what has been collected on daily basis*.” These testimonies support the interpretation that data cleanliness was treated not only as a statistical requirement but also as a routine managerial practice, although its depth varied across facilities.

### Descriptive findings

Descriptive statistics showed generally favourable but not uniformly strong perceptions across the five main explanatory constructs. Mean scores ranged from 3.09 for M&E capacity, 3.41 for M&E planning, 3.51 for M&E data management and use, and 3.45 for stakeholder perceptions of the DHFF M&E framework, to 3.73 for urban-rural M&E practice context. The dependent variable, DHFF implementation effectiveness, had an overall mean of 3.71, while organizational changes, behavioural changes and outputs scored 3.76, 3.95 and 3.83, respectively, suggesting that respondents perceived relatively stronger downstream implementation effects than foundational capacity conditions *(Table 7: Summary statistics – Mean scores for main variables)*.

**Table 7:** Summary statistics – Mean scores for main variables.

| Variable | Observations | Mean scores | Std. dev. | Min | Max |
| --- | --- | --- | --- | --- | --- |
| Quality of Health services | 233 | 3.714163 | 0.7104008 | 1.1 | 5 |
| Organizational changes | 233 | 3.759657 | 0.7506342 | 1 | 5 |
| Behavioural changes | 233 | 3.954506 | 0.8645254 | 1 | 5 |
| Outputs | 233 | 3.828326 | 0.7276351 | 1 | 5 |
| Mecap(CME) | 233 | 3.0897 | 0.7523391 | 1 | 4.5 |
| meplan(PME) | 233 | 3.414592 | 0.6802965 | 1.3 | 5 |
| medata(CASUME) | 233 | 3.514592 | 0.6872308 | 1 | 5 |
| meperce(SPMEF) | 233 | 3.454506 | 0.8307085 | 1 | 4.9 |
| mecontext(MEPERUC) | 233 | 3.729614 | 0.8007692 | 1 | 5 |
|  |  | Score range | Grading | Score coding |  |
|  |  | 1-1.8 | Strongly disagree | 1 |  |
|  |  | 1.9 -2.6 | Disagree | 2 |  |
|  |  | 2.7-3.4 | Sowhow agree | 3 |  |
|  |  | 3.5-4.2 | Agree | 4 |  |
|  |  | 4.3-5 | Strongly agree | 5 |  |

Disaggregated summaries sharpened this pattern. Within M&E data management and use, data utilization had the highest mean score (3.65), followed by sharing (3.52), collection (3.47) and analysis (3.41), indicating that respondents perceived data use forums and practical consumption of data somewhat more positively than analytical depth. Within stakeholder perception, effectiveness of the DHFF M&E framework (3.63) scored higher than its perceived relevance (3.27), suggesting that respondents observed practical gains even where conceptual ownership or fit remained weaker (*Table 8: Disaggregated summaries for sub variables*).

**Table 8:** Disaggregated summaries for sub variables.

| Variable | Observations | Mean scores | Std. dev. | Min | Max |
| --- | --- | --- | --- | --- | --- |
| meanqua l(DPIE) | 233 | 3.714163 | 0.7104008 | 1.1 | 5 |
| meanhring (Hrs) | 233 | 3.060086 | 0.8675143 | 1 | 5 |
| meantrain (Trn) | 233 | 3.124034 | 0.8271404 | 1 | 4.7 |
| meanplan (Pln) | 233 | 3.472532 | 0.7266604 | 1 | 5 |
| meanbudg (Bdt) | 233 | 3.342918 | 0.7436247 | 1 | 5 |
| meandcoll (Cln) | 233 | 3.474678 | 0.7404001 | 1 | 4.9 |
| meandanalys (Anly) | 233 | 3.414163 | 0.7882304 | 1 | 5 |
| meandshar (Shr) | 233 | 3.516309 | 0.6736354 | 1 | 5 |
| meanduse (Utl) | 233 | 3.64764 | 0.8385672 | 1 | 5 |
| meandrelev (Rlv) | 233 | 3.271674 | 1.000373 | 1 | 5 |
| meaneff (Eft) | 233 | 3.628755 | 0.8316408 | 1 | 5 |
| meanatt (Cpt) | 233 | 3.497854 | 0.9220688 | 1 | 5 |
| meanremot (Gre) | 233 | 3.729185 | 0.912607 | 1 | 5 |
| meanses (Scs) | 233 | 3.793991 | 0.8811033 | 1 | 5 |
| meandemand(Dms) | 233 | 3.774678 | 0.8732633 | 1 | 5 |
| meanutilcat (Hltu) | 233 | 3.785408 | 1.032386 | 1 | 5 |

Descriptive variation by level and setting also mattered. RHMT and CHMT respondents generally expressed stronger agreement across variables, whereas health centres and especially dispensaries showed more mixed responses, with a larger share of “somehow agree” answers and more visible implementation challenges. Differences are important because they suggest that the experience of DHFF-linked M&E becomes less robust as implementation shifts from supervisory levels to frontline facilities.

### Analytical findings

The aggregated regression model showed that the five main explanatory variables jointly had a statistically significant effect on DHFF implementation effectiveness, explaining about 52.4% of the variation in the outcome. In this model, M&E data management and use, stakeholder perceptions of the DHFF M&E framework, and urban-rural context emerged as the strongest significant contributors, whereas M&E capacity and M&E planning were positive but not statistically significant (*Table 9*). ANOVA confirmed the same pattern, with the model remaining significant and explaining about 56.8% of outcome variation, and again identifying data management and use, stakeholder perception and contextual practice as the most influential main predictors (*Table 10*).

**Table 9:**
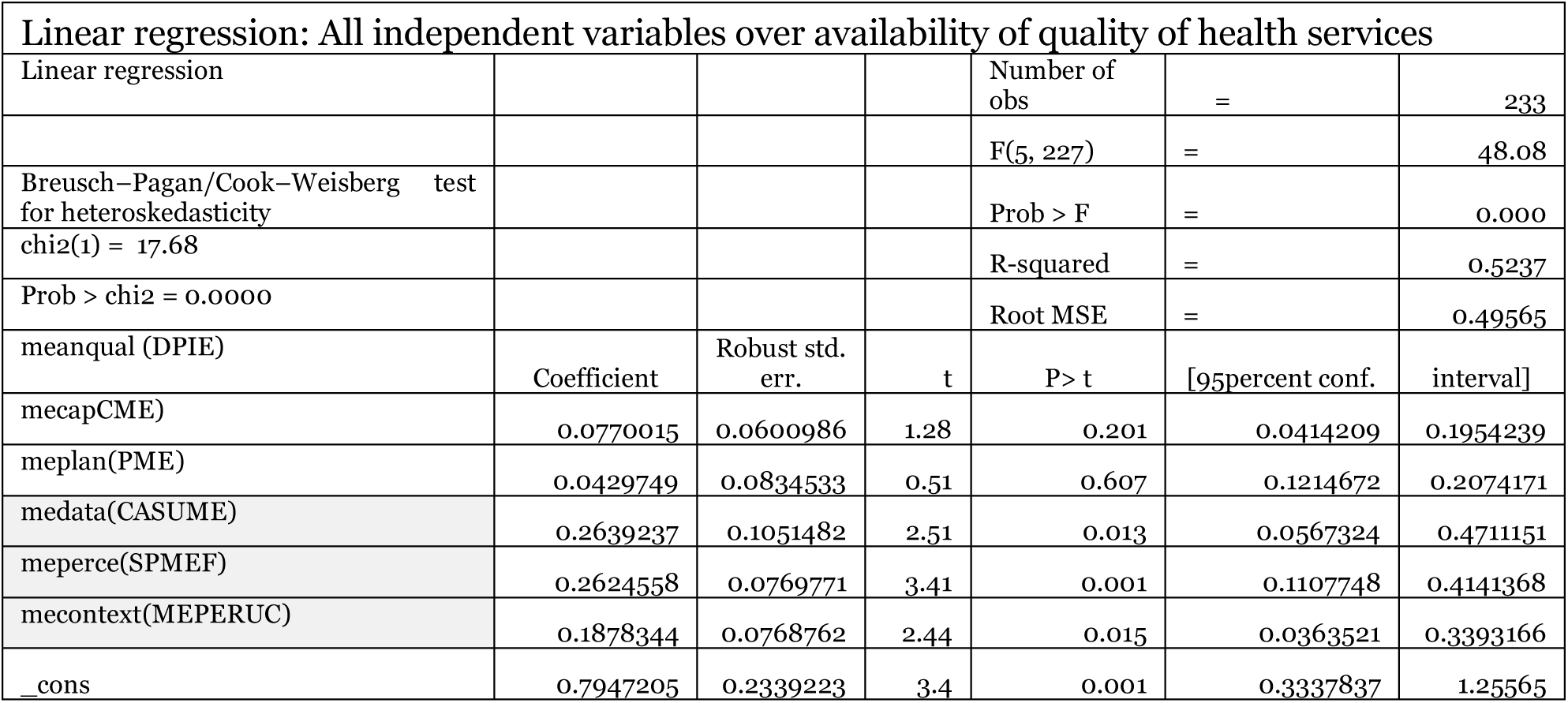
Overall regression model.

**Table 10:**
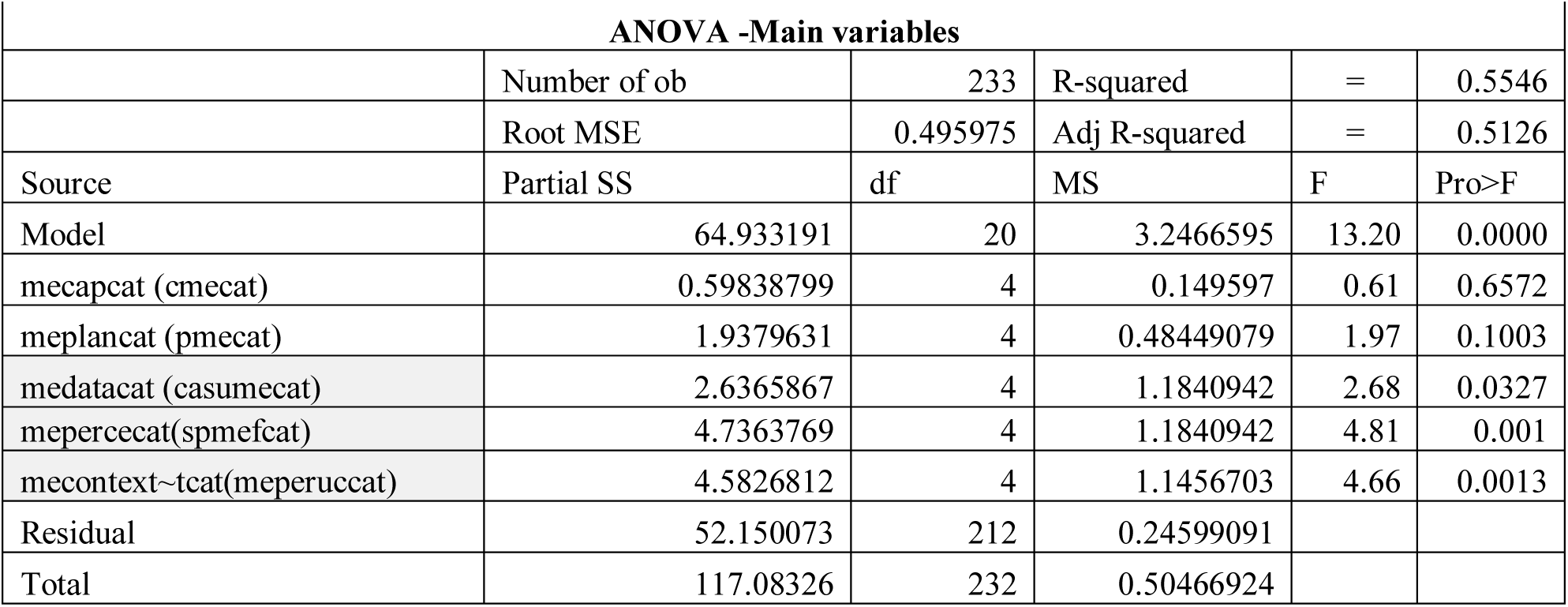
Overall ANOVA model.

The disaggregated analysis provided additional precision. Regression of the sub-variables explained 42.13% of variance in perceived quality health services, while the corresponding ANOVA showed adjusted explanatory power around 42.35%. Within the M&E data management domain, data utilization was the strongest significant sub-variable (p=0.018 in regression; p=0.0017 in ANOVA), and data analysis also showed meaningful influence, reaching marginal significance in regression (p=0.051) and significance in ANOVA (p=0.0269). By contrast, data collection and data sharing remained positive but statistically weaker in the multivariable models (*Table 11*).

**Table 11:** Regression - Availability of quality health services by M&E data management and use sub variables.

| Regression of availability of quality health services (DPIE) by M&E data management and use (CASUME) sub variables |  |  |  |  |  |  |
| --- | --- | --- | --- | --- | --- | --- |
| Number of obs |  |  |  |  |  | 233 |
| R-squared |  |  |  |  |  | 0.4213 |
|  |  | Robust |  |  |  |  |
| meanqual (DPIE) | Coefficient | std. err. | t | P>t | [95percent conf. | interval] |
| meandcoll (Cln) | 0.1132887 | 0.0838767 | 1.35 | 0.178 | 0.0519838 | 0.2785613 |
| meandanalys (Anly) | 0.1669602 | 0.08526 | 1.96 | 0.051 | 0.0010381 | 0.3349585 |
| meandshar (Shr) | 0.1596319 | 0.1216136 | 1.31 | 0.191 | 0.0799984 | 0.3992622 |
| meanduse (Utl) | 0.238021 | 0.0997157 | 2.39 | 0.018 | 0.0415388 | 0.4345031 |
| _cons | 1.320277 | 0.2267265 | 5.82 | 0 | 0.8735302 | 1.767024 |

Broader sub-variable modelling showed that perceived relevance of the DHFF M&E framework and health service utilization were the most consistent significant predictors of the outcome across the fifteen sub-components. The disaggregated ANOVA further indicated that M&E planning, data utilization, framework relevance, and geography/remoteness were especially influential in explaining outcome variation. Taken together, these analytical findings suggest that what matters most is not merely the presence of an M&E architecture, but whether data are used, interpreted as relevant, and applied within supportive operational contexts (*Table 12)*.

**Table 12:** ANOVA- Availability of quality health services by M&E data management and use sub variables.

| ANOVA of availability of quality health services (DPIE) by M&E data management and use (CASUME) sub variables |  |  |  |  |  |
| --- | --- | --- | --- | --- | --- |
| Number of obs | 233 |  |  | R-squared | 0.4632 |
|  |  |  |  | Adj R-squared | 0.4235 |
| Source | Partial SS | df | MS | F | Prob>F |
| meandcoll~t (Cln) | 2.2295298 | 4 | 0.55738244 | 1.92 | 0.1089 |
| meandanal~t (Anly) | 3.2602208 | 4 | 0.81505521 | 2.8 | 0.0269 |
| meandshar~t (Shr) | 1.7115347 | 4 | 0.42788368 | 1.47 | 0.2122 |
| meandusecat (Utl) | 5.1916363 | 4 | 1.2979091 | 4.46 | 0.0017 |
| Residual | 62.848326 | 216 | 0.29096447 |  |  |
| Total | 117.08326 | 232 | 0.50466924 |  |  |

### Integrating testimonies

Qualitative testimonies helped explain why data utilization outperformed other sub-variables analytically. One CHMT respondent from MDC stated, “*We have DHIS2 which is mother HIS being assisted by GoTHOMIS… we have FFARS… also PlanRep*,” showing the breadth of systems supporting data availability. Yet a facility respondent in KMC acknowledged a more limited analytic practice: “*We only add up numbers for the monthly report we don’t really analyze what they mean*.” This contrast mirrors the quantitative result that utilization and analysis not mere collection best distinguished stronger implementation outcomes.

The testimonies also clarified why lower-level facilities lagged behind. A facility respondent in MDC noted, “*Almost all staff or 99percent of staff at the facility have not been trained in IT and accountancy*,” while another explained, “*When we have technical problems, it can take two to three weeks to get help since there is only one IT personnel at council level for all facilities*.” Such statements illuminate the descriptive and inferential pattern in which rural and frontline sites reported lower performance on data analysis, timeliness and effective use.

Stakeholder-perception results were similarly reinforced by quotations. Some participants valued the framework’s transparency and autonomy, but HFGC members repeatedly described exclusion from data interpretation: “*We hear about DHIS2 and GoTHOMIS, but as HFGC we don’t see how they work or how to use them,*” and “*Sometimes the data feels like a mystery because we are told numbers but cannot verify them.”* These views align with the finding that framework effectiveness was rated higher than framework relevance, suggesting visible benefits without fully shared ownership.

Urban-rural contextual differences were also sharply reflected in testimonies. In KMC, a respondent stated, “*We use part of our cost-sharing funds to buy internet bundles and print reports when needed,*” while in MDC a CHMT respondent explained, “*Internet connectivity is a very big problem, which disturbs the equilibrium of data flow at required times in all health facilities*.” These testimonies provide connective evidence for the statistical result that contextualized M&E practice significantly shaped DHFF implementation effectiveness.

## Discussion

This study shows that M&E data management and use are among the strongest determinants of DHFF implementation effectiveness, and that this effect persists even after accounting for stakeholder perceptions and contextual differences between urban and rural settings. This finding is consistent with Tanzanian evidence showing that data use, not data availability alone, is what links routine information systems to planning, budgeting, accountability, and service improvement under decentralized health financing arrangements (5–10,33,42,44,61). It is also consistent with broader African and global literature showing that interoperable systems, digital tools, and reporting platforms create value only when they are embedded in governance, feedback, and decision-use processes (11–23,30,40,45,47–53,65).

The prominence of data utilization and data analysis over data collection and data sharing in the present study is especially important. It suggests that implementation value is created less by producing reports alone than by interpreting findings, discussing them in management forums, and applying them to local decisions, which echoes prior work from Tanzania on council planning, facility-level HMIS use, and the institutionalization of routine data use at sub-national level (6–10,44,61). Related evidence from Ethiopia, Uganda, Mali, and wider digital-health literature similarly shows that data quality, timeliness, interoperability, and analytic culture are critical if routine systems are to support meaningful corrective action rather than administrative compliance only (12–15,18–22,40,45,47–49).

The descriptive findings add important nuance to this interpretation. Although respondents generally rated all major constructs positively, M&E capacity remained the weakest foundational condition, while context-sensitive M&E practice scored highest, suggesting that implementation strength is shaped not only by formal systems but also by local operational realities. This reading aligns with Tanzanian and regional evidence showing that health-system performance depends on how governance, staffing, infrastructure, supervision, and decentralization function in practice rather than on formal policy architecture alone (24,28,31,35,36,50,51,56,57,59,64).

The qualitative evidence deepens this explanation by showing how local actors distinguish between having digital platforms and being able to use them meaningfully. The coexistence of DHIS2, GoTHOMIS, FFARS, PlanRep, eLMIS, and NEST reflects Tanzania’s wider investments in digitalization and interoperability, yet the testimony that some facilities mainly “add up numbers” for monthly reports illustrates the persistent gap between system presence and analytic use (5,18,19,21,22,33,40,42,45,47–49,53). This pattern is increasingly recognized in recent digital-health analyses, which argue that digital transformation succeeds only when tools are matched to context, user capability, governance arrangements, and implementation support (19,21,22,40,45,47–49,53).

Stakeholder perceptions of the DHFF M&E framework emerged as another central explanatory pathway. This is consistent with literature on participatory monitoring, social accountability, health-facility governing committees, and decentralized planning, which suggests that actors are more likely to use data when they regard monitoring arrangements as relevant, understandable, inclusive, and actionable (16,17,23,24,28,29,37,50,51,60). The gap observed in this study between perceived framework effectiveness and perceived relevance therefore matters: it suggests that a framework may produce administrative outputs and some service gains even when community actors and frontline implementers do not feel genuine ownership of the monitoring process (2,24,27,37,50,60).

Urban-rural context also clearly conditioned the effect of M&E on DHFF implementation. This finding is supported by evidence that rural facilities in Tanzania and comparable settings face greater constraints in connectivity, staffing, service readiness, financial protection, and timely technical support, all of which reduce the practical utility of data systems and weaken feedback loops (15,24,25,32,35–37,41,54,56,57). By contrast, better-resourced urban settings are more likely to sustain routine review meetings, internet access, printing, troubleshooting, and cross-team coordination, thereby allowing the same formal M&E architecture to function more effectively in practice (5–10,24,32,36,42,44,56,57,61).

Another important implication is that implementation bottlenecks are concentrated at the point where reporting burden and service-delivery pressure intersect. The weaker experiences reported at health-centre and dispensary level fit with prior evidence that frontline facilities often have fewer highly trained staff, less analytic support, and greater competition between clinical work and reporting responsibilities (6–10,11,13,14,30,44,55,61). This means that strengthening DHFF implementation will require attention not only to data systems themselves, but also to workload organization, mentoring, staffing distribution, and practical visualization tools that make local data easier to interpret and use (11,18,19,22,30,43–45).

The broader interpretation of the outcome measures is that DHFF appears to be generating visible gains in organizational discipline, accountability, service readiness, and selected output-level improvements even where formal M&E capacity remains moderate. This is in line with evidence that DHFF and related provider-autonomy reforms can improve budget execution, local responsiveness, and some aspects of quality or maternal service delivery, while still leaving unresolved gaps in analytic depth, equity, and participatory governance (1–4,27,38,39,57,62,63). In practical terms, the findings suggest that the durability and equity of DHFF gains will depend on whether data-use systems become more inclusive, more context-responsive, and more closely tied to local problem-solving.

From a policy standpoint, the combined descriptive, analytical, and qualitative evidence points to three priorities. First, capacity-building should focus less on reporting compliance alone and more on interpretation, visualization, feedback, and decision use, especially at health-centre, dispensary, and HFGC levels (11–14,18,19,22,30,43–45). Second, rural facilities require targeted digital and supervisory support, including faster troubleshooting, offline-capable workflows, and more equitable distribution of technical assistance and infrastructure (6–10,15,21,32,35,40,42,45,47,51–57). Third, the DHFF M&E framework needs stronger participatory translation so that HFGCs and community actors do not remain passive recipients of figures but become effective users of data in governance, accountability, and local priority-setting (16,17,23,24,28,29,37,50,51,60).

### Limitations

This article is based on findings from two councils and therefore reflects deep contextual insight rather than national representativeness. In addition, some measures of implementation effectiveness and M&E practice rely on stakeholder perceptions, which may be influenced by recall or social desirability bias. Nevertheless, the use of mixed methods, triangulation and comparison across urban and rural settings strengthens the credibility of the interpretations presented here.

## Conclusion

M&E data management and use have a substantial positive effect on DHFF implementation effectiveness in Tanzania, but this effect is mediated by stakeholder perceptions of the DHFF M&E framework and by urban–rural contextual conditions. Stronger data systems alone are not enough; what matters is whether facilities and governance actors can routinely interpret and use data within an enabling environment of skills, infrastructure, communication and accountability. Sustaining DHFF gains will therefore require integrated investments in data use capacity, digital support, participatory governance and context-sensitive implementation strategies that close persistent rural disadvantages.

## Author summary

This article examined how monitoring and evaluation data management and use influence the effectiveness of Direct Health Facility Financing (DHFF) in Tanzania. Drawing on findings from an urban council and a rural council, it shows that facilities that routinely use data for planning, accountability and service improvement tend to perform better under DHFF than those that mainly report data for compliance. The study also shows that stakeholder perceptions of the DHFF M&E framework and the differences between urban and rural settings help explain why data are used more effectively in some facilities than in others. The results suggest that improving DHFF implementation requires not only better data systems, but also stronger training, participation, infrastructure and feedback mechanisms, especially in rural areas.

## Data availability

All data used to develop this article were derived from the study thesis and can accessed when requested from author

## Author contributions

**Conceptualization:** Deogratias F Mpenzi.

**Methodology:** Deogratias F Mpenzi; Deus D Ngaruko; Roger Myrick.

**Formal analysis:** Deogratias F Mpenzi.

**Investigation:** Deogratias F Mpenzi, Deus D Ngaruko; Roger Myrick.

**Writing – original draft:** Deogratias F Mpenzi.

**Writing – review & editing:** Deogratias F Mpenzi, Deus D Ngaruko; Roger Myrick.

**Supervision:** Deus D Ngaruko; Roger Myrick.

**Validation:** Deus D Ngaruko; Roger Myrick.

## Data Availability

Minimal data set shall be made available when required during the review process. The authors developed code book and prepared a data set in the excel format and captured data from the data collection tool, then uploaded it into STATA version 18 for cleaniliness and processing for descriptive analyis and inferential analysis
so all data produced in the present study are available upon reasonable request to the authors

